# A Novel Therapy for ALS: Allogeneic Schwann Cell Extracellular Vesicles

**DOI:** 10.1101/2023.01.18.23284378

**Authors:** Pascal J. Goldschmidt-Clermont, Aisha Khan, James D. Guest, George Jimsheleishvili, Patricia Graham, Adriana Brooks, Risset Silvera, Alexander J.P. Goldschmidt, Damien D. Pearse, W. Dalton Dietrich, Robert Brown, Allan D. Levi

## Abstract

Amyotrophic Lateral Sclerosis (ALS) is a terminal condition with accelerated loss of motor neurons (MN), resulting in the progressive paralysis of affected patients. ALS is either sporadic (90%) or genetically transmitted (10%) and affects cortical (pyramidal) and spinal cord (lower) MN, axons, and respective muscle endplates. ALS research has focused on MN survival, and current FDA-approved therapies provide only small patient survival benefits. This study reports the intravenous (IV) delivery of serial infusions of allogenic Schwann cell-derived extracellular vesicles (SCEV). The recipient had transient clinical stabilization during treatment but deteriorated rapidly during a pause in the infusions. There were no SCEV infusion-related adverse events observed. Allogeneic SCEV appeared safe for IV delivery in this case and may have therapeutic potential.

## Case report

Previously, in spinal cord injury (SCI) studies, we assessed the ability of transplanted autologous human Schwann cells (ahSCs) to improve neurological function. We have completed a series of FDA-approved phase I clinical trials in which culture-expanded ahSC were injected into the spinal cord lesion of paralyzed victims with sub-acute or chronic SCI^1,2^. The therapy appears safe and is being advanced to increase the impact on the repair of spinal cord and nerve injuries^3^. Schwann cells (SC) are essential for the maintenance and function of MN, and their function may be impaired in ALS^4-6^. Recently, important signaling and potential trophic functions of exosomes released by SC have been reported^7-9^.

We report a case of a man in his 8^th^ decade with a 1.5-year history of progressive ALS. After initial diagnosis, the patient was ambulatory at home for four months, became wheelchair-bound after eight months, required automatic positive airway pressure (APAP) respiratory support after 12 months, and became APAP machine-dependent 24/7 after 14 months. The patient volunteered to participate in an FDA-approved single-patient expanded access treatment for ALS, delivering SCEV weekly intravenously to rescue impaired SC and MN function.

## Methods

SC were extracted from a nerve of a cadaveric donor. Screening of the cadaver allogeneic donor met eligibility criteria as outlined in 21 CFR Part 1271. SC were dissociated from the nerve and culture-expanded, as previously reported^10^. SCEV were collected from culture media and tested negative for sterility and mycoplasma; endotoxin was <0.5EU/kg body weight. The particle concentration was 3.74e+11 +/- 9.28e+09 with a modal size of 137.0 nm. The protein content was 0.48 mg/ml, and CD81 and CD 63 expression were above 90% (refer to Table 1).

**Table 1.** Release Testing of SCEV

| Sample | Assay | Results |
| --- | --- | --- |
| SCEVs | Sterility | Negative |
|  | Endotoxin | <0.5EU/kg body weight |
|  | Mycoplasma | Negative |
|  | Particle Concentration | 3.74e+11 +/- 9.28e+09 |
|  | Particle Mode size: between 50 and 200 nm | 137.0 |
|  | Protein Content | 0.48 mg/ml |
|  | Characterization by Flow Cytometry Analysis | CD81 – 90.5 %<br>CD63 – 95.5% |

Each week before SCEV infusions, in addition to a complete physical examination, the patient was evaluated with the ALS Functional Rating Scale-Revised (ALSFRS-R), and for pulmonary function with a spirometry device [Medical Technologies, EasyOne Air] administered by a trained assessor under consistent conditions. Refer to Tables 2 and 3 for the patient’s assessment schedule.

**Table 2.** Time and Events Schedule for 2 low and 2 medium doses.

|  | Baseline | Day 0 | Day 1 | Day 2 | Day 7 (+/- 2 days) | Day 8 | Day 9 | Day 14 (+/- 2 days) | Day 15 | Day 16 | Day 21 (+/- 2 days) | Day 22 | Day 23 |
| --- | --- | --- | --- | --- | --- | --- | --- | --- | --- | --- | --- | --- | --- |
| Demographics | X |  |  |  |  |  |  |  |  |  |  |  |  |
| Medical History | X |  |  |  |  |  |  |  |  |  |  |  |  |
| Physical Exam | X | X |  |  | X |  |  | X |  |  | X |  |  |
| Vitals<sup>a</sup> | X | X |  |  | X |  |  | X |  |  | X |  |  |
| Telephone Call |  |  | X | X |  | X | X |  | X | X | X | X | X |
| CBC and CMP | X | X |  |  | X |  |  | X |  |  | X |  |  |
| ALSFRS-R<sup>b</sup> | X |  |  |  | X |  |  | X |  |  | X |  |  |
| Spirometry | X |  |  |  | X |  |  | X |  |  | X |  |  |
| **SCEVs IV Infusion** |  | **X** |  |  | **X** |  |  | **X** |  |  | **X** |  |  |
| Biomarkers:<br>• CRP<br>• Serum NFL<sup>c</sup><br>• IL6, IL8, IL5 & IL2 | X | X |  |  | X |  |  | X |  |  | X |  |  |
| Adverse Event Monitoring | X |  |  |  |  |  |  |  |  |  |  |  |  |

<sup>a</sup> Vitals will be done at specific intervals pre, during and post infusion: refer to section on **Infusion Guidelines & Post Infusion Monitoring**<sup>b</sup> Amyotrophic Lateral Sclerosis Revised Functional Rating Scale (ALSFRS-R)<sup>c</sup> Neurofilament Light Chain (NFL)**Table 3.** Time and Events Schedule for 4 medium doses.

**Table 3.** Time and Events Schedule for 4 medium doses.

|  | Day 28 (+/- 2 days) | Day 29 | Day 30 | Day 35 (+/- 2 days) | Day 36 | Day 37 | Day 42 (+/- 2 days) | Day 43 | Day 44 | Day 49 (+/- 2 days) | Day 50 | Day 51 |
| --- | --- | --- | --- | --- | --- | --- | --- | --- | --- | --- | --- | --- |
| Demographics |  |  |  |  |  |  |  |  |  |  |  |  |
| Medical History |  |  |  |  |  |  |  |  |  |  |  |  |
| Physical Exam | X |  |  | X |  |  | X |  |  | X |  |  |
| Vitals<sup>a</sup> | X |  |  | X |  |  | X |  |  | X |  |  |
| Telephone Call |  | X | X |  | X | X |  | X | X | X | X | X |
| CBC and CMP | X |  |  | X |  |  | X |  |  | X |  |  |
| ALSFRS-R<sup>b</sup> | X |  |  | X |  |  | X |  |  | X |  |  |
| Spirometry | X |  |  | X |  |  | X |  |  | X |  |  |
| **SCEVs IV Infusion** | **X** |  |  | **X** |  |  | **X** |  |  | **X** |  |  |
| Biomarkers:<br>• CRP<br>• Serum NFL<sup>c</sup><br>• IL6, IL8, IL5 & IL2 | X |  |  | X |  |  | X |  |  | X |  |  |
| Adverse Event Monitoring | X |  |  |  |  |  |  |  |  |  |  |  |

<sup>a</sup> Vitals will be done at specific intervals pre, during and post infusion: refer to section on **Infusion Guidelines & Post Infusion Monitoring**<sup>b</sup> Amyotrophic Lateral Sclerosis Revised Functional Rating Scale (ALSFRS-R)<sup>c</sup> Neurofilament Light Chain (NFL)

## Results

To determine if SC dysfunction could contribute to our patient’s ALS, we obtained a biopsy of his left sural nerve (5 cm) under local anesthesia. We placed it in culture in an optimized SC medium to expand and study the patient’s SC^10^. His SC appeared senescent and proliferated poorly (Figure 1A), and the SC were contaminated with fibroblasts (Figure 1B). When cultured with allogeneic control SCEV generated from non-ALS donor SC, the patient’s SC had more robust SC marker expression (CD 271) at 38.51% (Figure 1D), as compared to 20.43% (Figure 1C) without the non-ALS SCEVs. In addition, fewer contaminating fibroblasts (CD90) were observed, 29.10% (Figure 1D) as compared to 50.65% (Figure 1C). This experiment suggests that the peripheral nerve SC of the patient were poorly functional, possibly contributing to declining MN function. Based on these findings, we hypothesized that the extracellular vesicles released by healthy control SC from an allogeneic donor might improve function in diseased ALS SC, MN, and possibly muscles.

**Figure 1.**
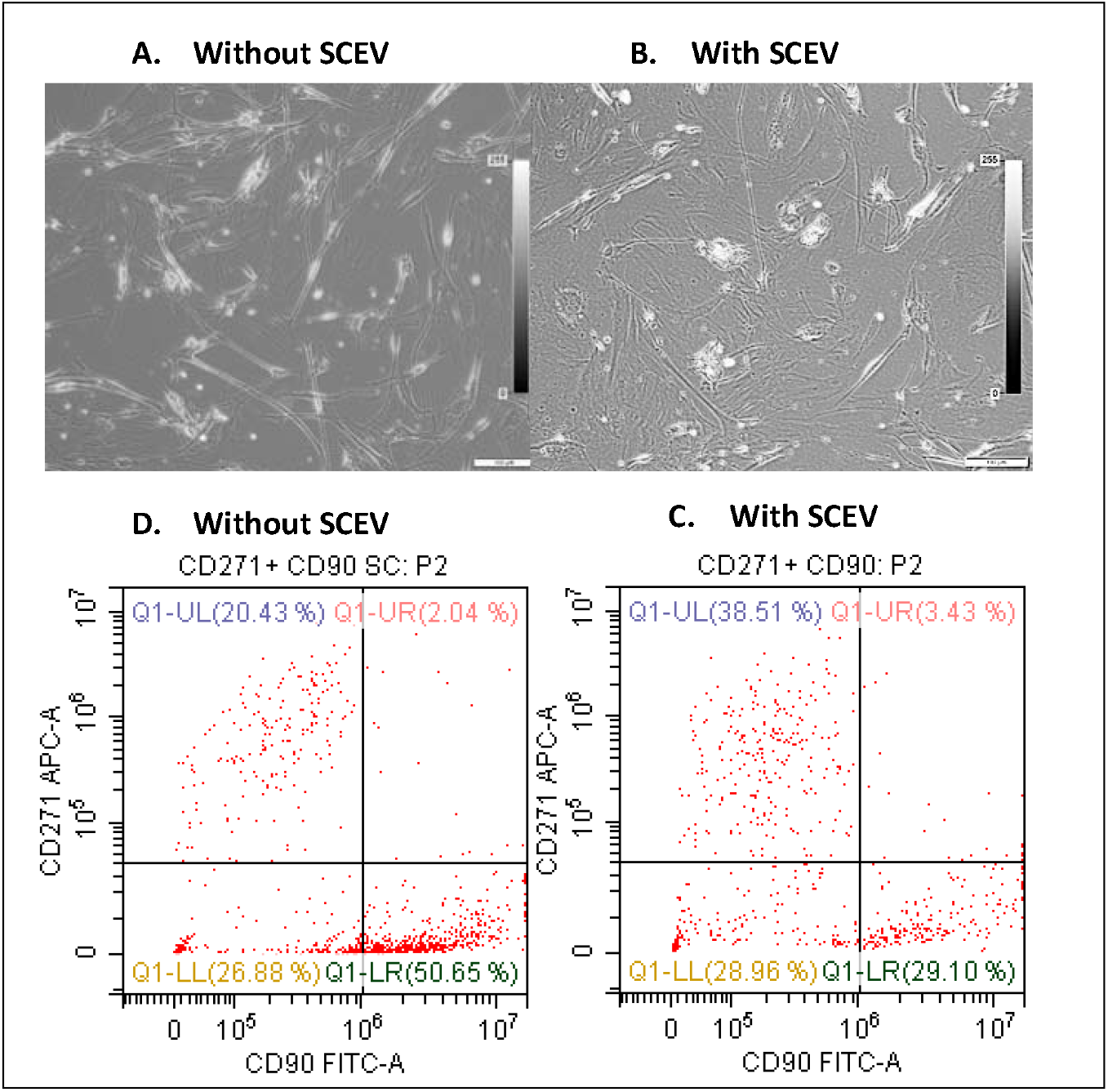
Schwann cell cultures from the subject with and without allogenic SCEV from a donor without ALS. Phase contrast microscope, 10x. **A**. Without, and **B**, with allogeneic SCEV, added. **Panels C** and **D**. Flow cytometric analysis for Schwann cell marker CD271, and fibroblast marker CD90, same conditions as **A** and **B**. In **D**, with allogeneic SCEV, the percent of CD271 is increased, and CD90 is decreased.

To test this idea, we designed a protocol of weekly allogeneic SCEV infusions for our patient. The first infusion containing 1.54 × 10^12^ SCEV diluted in 40ml of DPBS was accomplished without allergic reaction or adverse effects related to SCEVs in the Intensive Care Unit, University of Miami Hospital. After that, for the patient’s comfort, subsequent infusions were administered at his home with FDA’s approval. A dose escalation occurred for the third infusion (7.5 × 10^12^ SCEV diluted in 40ml of DPBS). Subsequently, five more infusions (7.5 × 10^12^ SCEV) were delivered. Thus, the patient received eight weekly infusions, two low doses, and six medium doses. Due to a lack of ability to generate sufficient weekly SCEV, the treatment was temporarily paused. None of the infusions were associated with adverse effects such as infusion reactions or changes in vital signs. The high dose (15 × 10^12^) of SCEV was supposed to start in January. The investigators were surprised by the rebound of the disease process after a short interruption of the infusions. If supported by future studies, this therapy may require long-term administration of SCEV.

Six standard measurements were performed ten weeks before infusions started to document the ALS baseline progression, then nine times over the subsequent nine weeks to assess the impact of SCEV infusions. The only significant patient complaint was difficulty in expectorating phlegm about once a day, a known ALS complication that continued after the initiation of infusions. Complete blood work (see additional online information) was obtained the week before the first infusion, then each week just before initiating the infusion. No notable changes in the cytokines IL-6, IL-10, or the biomarker neurofilament light chain were observed.

The pre-infusion rate of functional decline (ALSFRS-R)^11^ shown in Figure 2 averaged 1.71 points/month, followed by seven measurements during the infusions period with an average rate of decrease of 1.22 points per month (difference 0.42 points per month). Then the last two measurements were obtained after the discontinuation of the infusions. The shaded boxes (Figure 2) illustrate an apparent stabilization of the disease during the infusion period.

**Figure 2.**
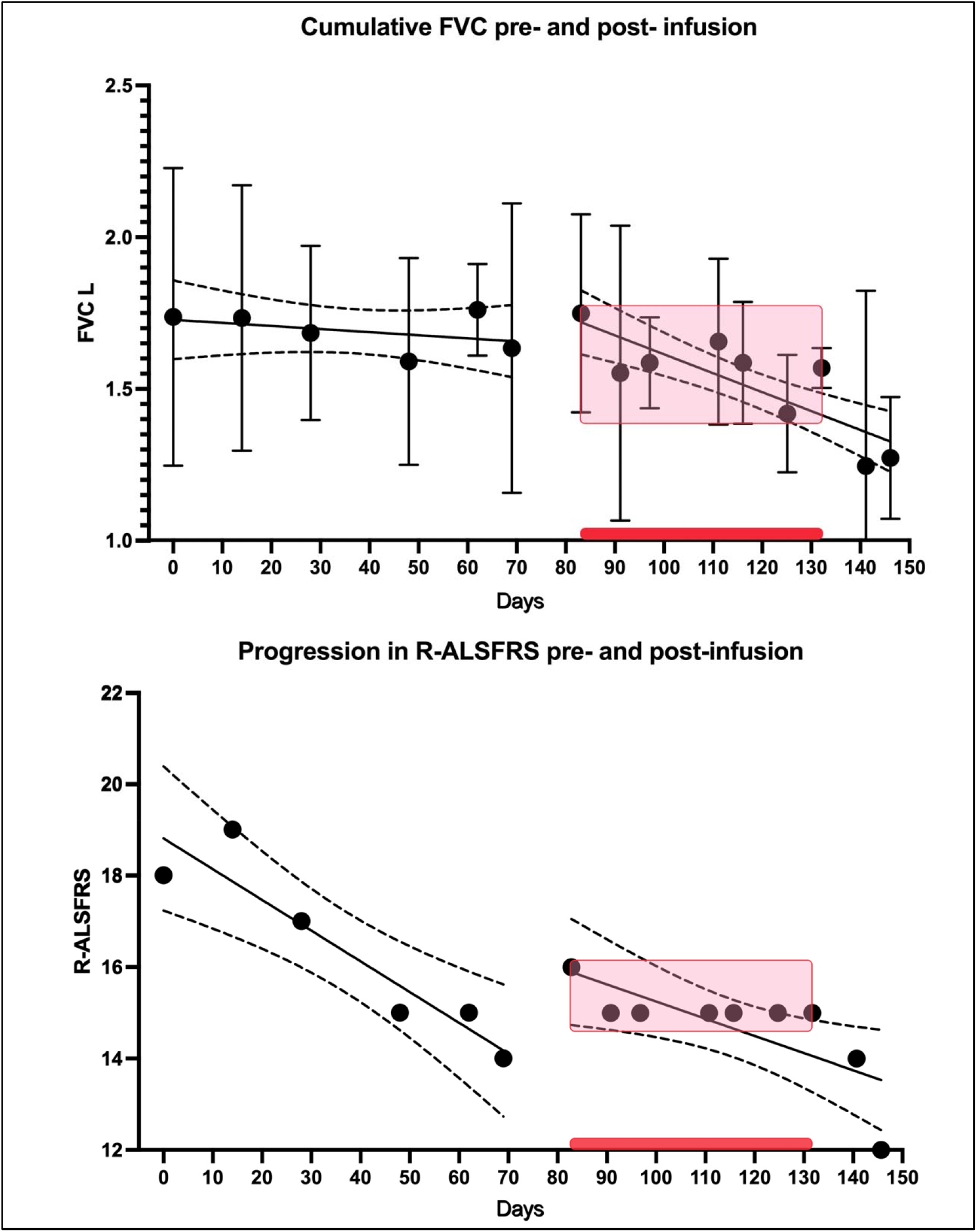
Values of Forced Vital Capacity and ALSFRS-R during the study. The pre-infusion period extended until 70 days. The period in which the seven infusions occurred is indicated on the x-axis with the red bar. There were two values after the infusions were paused. The stabilization of values is indicated by the pink boxes. Overall slopes are shown for the pre-infusion and infusion periods, including all values. FVC in liters.

Pre-infusion pulmonary function declined as expected in ALS^12^ (forced vital capacity in liters measured by spirometry), as shown in Figure 2 lower panel. The following seven measurements correspond to the infusions period, and the last two measurements were obtained after the discontinuation of the infusions.

While no significant adverse events related to infusions were noted during the entire infusion period, two days after the last infusion, the patient complained of diarrhea temporally associated with an elevation of c-reactive protein (CRP, from 2 to 20 mg/dl). Then a week later, two days after blood work that showed decreased CRP and no elevation of his white blood cells and neutrophils, the patient presented with acute respiratory destabilization and was admitted to the hospital and needed emergent endotracheal intubation and ventilation (the patient had requested to remain full code). In addition, stress indices were highly elevated, neutrophil-lymphocytes ratio^13,14^ of 39.8 and a platelet lymphocyte ratio of 536.

His workup indicated the presence of right upper lobe pneumonia on X-ray and significant WBC elevation with neutrophilia. The patient requested the removal of his endotracheal tube and rejected the offer of advanced respiratory care, including a tracheostomy. He was switched to comfort care and passed away.

## Discussion

ALS is an incurable condition where the patients remain fully aware of their decline as cognitive functions are relatively preserved^15^. Most patients die within three years of their diagnosis^16^. During this study, our patient received state-of-the-art treatment for ALS with two FDA-approved drugs approved by the FDA, riluzole 50mg twice daily^17^, and Relyvrio (a combination of sodium phenylbutyrate, 6 grams per day, and tauroursodeoxycholic acid 1gram per day)^18^. Notably, the reduction in ALSFRS-R during SCEV infusions compared to before infusions (0.49 points/month) is similar to the average benefit reported in the CENTAUR study of Relyvrio™ (0.42 points/month)^18,19^.

A notable finding, if reproducible with multiple ALS patients, is that the peripheral nerve SC of the participant were poorly functional, possibly contributing to the decline of MN function, but showed partial culture rescue with allogeneic non-ALS SCEV. Our study provides a novel approach to address impaired SC and MN function in ALS using allogenic SCEV. Multiple infusions were not associated with adverse reactions, and there was a period of disease stabilization. Further exploratory studies are warranted.

## Data Availability

CARE guideline followed.

## Acknowledgment

The study investigators are grateful to Michael Benatar, MD, Ph.D., for his ALS expertise provided during the study, for the invaluable nursing skills of Rosie Igheldane, RN, BSN, and Sabrina Taldone, MD, for medical input. Financial support from the Miami Project to Cure Paralysis, the Buoniconti Fund, and the Interdisciplinary Stem Cell Institute, and the unconditional support of Dean Henri Ford of the Leonard M. Miller School of Medicine at the University of Miami.

